# Data Resource profile: Medicines in Acute and Chronic Care in Scotland (MACCS)

**DOI:** 10.64898/2026.03.19.26348795

**Authors:** Cosmika Goswami, Tanja Mueller, Amanj Kurdi, Ewan Pearson, Khaled Bedir, Alexandra Tolfrey, Hazel Close, Marion Bennie

**Affiliations:** Strathclyde Institute of Pharmacy and Biomedical Sciences, University of Strathclyde, UK; Public Health Scotland, Edinburgh, UK; College of Pharmacy, Al-Kitab University, Kirkuk 36015, Iraq; Department of Public Health Pharmacy and Management, School of Pharmacy, Sefako Makgatho Health Sciences University, Pretoria, South Africa; Faculty of Health, University of Dundee, UK

## Abstract

**Background:** Routinely collected prescribing and medicine-related data in Scotland are comprehensive and of high quality. However, they are generated across multiple healthcare settings and stored in complex source systems that are not optimised for longitudinal or outcomes-focused research. To maximise the research value of these data, there is a need for curated, analysis-ready resources that provide consistent representations of medicines exposure and enable linkage to clinical outcomes. The Medicines in Acute and Chronic care Scotland (MACCS) provides standardised, curated medicines data to support longitudinal analyses of medicine-related exposure across NHS healthcare systems.

**Methods:** MACCS resource is a national individual-level medicines dataset for adults (18 years of age and older), derived from routinely collected prescribing and medicine-related data held by Public Health Scotland (PHS). It integrates data from the Hospital Electronic Prescribing and Medicines Administration (HEPMA), Prescribing Information System (PIS), and Homecare Medicines (HCM) datasets, which are linked at the individual level to eleven other national clinical records; including Scottish Morbidity Records (SMR00/01/02/04/06), laboratory data and mortality records; using the unique NHS Scotland person identifier. Data are curated, harmonised and pre-linked within the National Safe Haven and accessed by approved researchers through secure Trusted Research Environments.

**Results:** MACCS contains individual-level information on adults receiving NHS Scotland care, including patient demographics (such as age, sex and geographical indicators) and detailed records of medicines prescribing in community pharmacies as well as those administered in hospitals and through homecare services. Medicines-related data captures exposure dates and, where available, details on formulation, strength and dose. In addition, MACCS includes cancer registry data, renal registry data, laboratory test results, microbiology surveillance and mortality records. The earliest dates of data availability vary by source dataset.

**Conclusion:** MACCS provides a sustainable, longitudinal medicines research resource that simplifies access to complex national prescribing data and enables robust linkage to health outcomes. By supporting population-scale analyses across care settings, MACCS enhances the capacity for high-quality research to inform clinical practice, health policy, and medicines optimisation in Scotland.

**Key Features:**

- The Medicines in Acute and Chronic Care in Scotland (MACCS) data resource was established in 2025 to integrate medicine-related data with other electronic data from Scottish healthcare systems, creating a national, linked, routinely updated data resource at population level.
- MACCS provides pre-linked data from multiple routinely collected national datasets within NHS Scotland including, but not limited to, prescribing records, hospital episodes, laboratory results, and death records, within a single secure environment.
- MACCS includes patient demographics, data on medicines prescribing and administration/supply, key biochemistry and haematology test results (e.g., kidney and liver function tests), data on hospital admissions and surgical procedures, and date and cause of death.
- The data resource provides longitudinal follow-up of the adult population (≥18 years of age) receiving medicines through NHS Scotland since 2010, covering approximately 4.6 million individuals, and supports pharmacoepidemiological studies, drug utilisation research, pharmacovigilance projects, as well as health services research.
- Approved researchers can apply through a streamlined process to access the linked MACCS data resource through established NHS Scotland governance processes, with data accessed within a Trusted Research Environment.

## Data resource basics

Healthcare in Scotland is delivered through a universal, publicly funded system operated by NHS Scotland, under which services are provided free at the point of care to all residents. This single-payer structure enables comprehensive population coverage and the routine collection of administrative health data at a national level [1–3]. NHS Scotland serves a population of approximately 5.3 million people [1], encompassing both urban and rural settings and a wide range of socioeconomic circumstances. Over recent years, electronic systems have been implemented to support the prescribing and dispensing/administration of medicines in both primary and secondary care [4, 5], to complement already existing electronic systems providing a wide range of functionalities across the healthcare system [6]. However, because medicines are prescribed in different healthcare settings, namely primary care, secondary care and home care, information is captured in separate datasets reflecting the context of care delivery. Furthermore, medicines-related data are stored separately from other routinely collected healthcare data such as hospital or death records due to data being collected through various, distinct electronic systems. While these electronic systems have strengthened clinical practice and patient safety within individual settings, the resulting fragmentation of data presents challenges for research, including attempts to understand patterns of medicines use and associated outcomes across the healthcare system as a whole [6–11]. The Medicines in Acute and Chronic Care in Scotland (MACCS) data resource was established in 2025 to address this gap by providing comprehensive, longitudinal data on medicines use across all healthcare settings in Scotland, pre-linked to a range of other healthcare datasets – including outpatient clinic attendances, in-patient hospital episodes, select laboratory test results, disease registries, and death records. Record linkage is facilitated by the availability of a unique patient identifier in Scotland, allocated to every resident and included in all health and social care records [12]. MACCS covers all adults (18 years of age or older) residing in Scotland who have received at least one prescription medicine since 2010; at the time of writing (February 2026), MACCS comprises data on approx. 4.6 million individuals.

MACCS is based on existing routinely collected data and data use is governed through established NHS Scotland information governance processes. Approval for the establishment of the MACCS data resource was obtained through the Public Benefit and Privacy Panel for Health and Social Care (PBPP-HSC) [13]; MACCS is held and maintained by Public Health Scotland (PHS) and is part of the PHS infrastructure, with initial development support provided by Health Data Research UK and the Scottish Government Pharmacy, Medicines and Therapeutics Division. MACCS is intended as a long-term, curated data resource contingent on the continued operation of national prescribing systems and public sector funding arrangements, to be updated annually, for use by approved researchers within a Trusted Research Environment [14].

## Data collected

MACCS is derived entirely from routinely collected administrative healthcare data and integrates information from multiple existing national datasets within NHS Scotland. Individuals are included in MACCS on the basis of medicine exposure: the eligibility criteria for inclusion are being aged 18 years or older; and having received at least one prescription medicine within NHS Scotland – dispensed through a community pharmacy since 04.2009; administered in hospital since 07.2022; and/or delivered directly to their home through the Medicines Home Care Services since 01.2019. Extensive clinical, demographic, and laboratory information is available for included individuals through record linkage via the Community Health index (CHI) number, a unique person identifier which enables deterministic linkage across the datasets [12]. Record linkage is performed by the Electronic Data Research and Innovation Service (eDRIS) [15] within PHS and follows established information governance procedures.

MACCS currently combines information from 14 different sources, thereby providing data on medicines use; medical conditions including laboratory data; hospitalisations; deaths; and socio-demographics. National medicines datasets include Prescription Information System (PIS), holding information on medicines supplied in community pharmacies [4]; Hospital Electronic Prescribing and Medicines Administration (HEPMA), covering inpatient wards in hospitals (excluding systemic anti-cancer therapy and intensive care units) [5]; and Home Care Medicines (HCM), i.e., data on medicines prescribed in secondary care and delivered directly to a patient’s home (typically highly specialist medicines such as biologics) [16]. Disease registries such as the Cancer Registry (SMR06)[17] and the Scottish Renal Registry (SRR)[18] provide data on medical conditions; key biochemistry and haematology test results (e.g., kidney and liver function tests) are held in the Scottish Care Information (SCI) Store [19] while microbiology surveillance data stem from the Electronic Communication of Surveillance Scotland (ECOSS) [20]. Data on hospitalisations is provided through Scottish Morbidity Records (SMR) covering outpatient, inpatient, maternity, and mental health care (SMR00/01/02/04) [21–24]; and the Scottish Intensive Care Society Audit Group (SICSAG) [19] covering critical care. Death records from the National Records of Scotland (NRS) are available to provide data on mortality [25]. Patient demographics – including age, sex, health board and urban-rural classification of residence, and the Scottish Index of Multiple Deprivation (SIMD) [26] are available across the datasets; see Table 1 for further details on the individual datasets. Work is in progress to add further data; in the first instance, Unscheduled Care Data comprising data from Emergency Departments, GP out-of-hours systems, and the Scottish Ambulance Service. Discussions are ongoing to also include primary care data (data from GP records).

**Table 1:** Data provided in the Medicines in Acute and Chronic Care in Scotland (MACCS) data source.

| Dataset | Dataset Description | Type of Data Collected | Data Coverage |
| --- | --- | --- | --- |
| **Medicines use data** |  |  |  |
| PIS [27] | Medicines prescribed and dispensed in primary care | Medicine name, formulation, strength; date prescribed, date dispensed; quantities prescribed and dispensed | 04.2009 – 02.2025 |
| HEPMA [5] | Medicines prescribed and administered in hospital | Medicine name, dose, route; date prescribed, date and time administered | 07.2022 – 05.2025 |
| HCM [28] | Medicines prescribed in secondary care and delivered directly to patients' homes | Medicine name, quantity, supply date, indication | 01.2019 – 08.2025 |
| **Clinical information** |  |  |  |
| SMR06 [17] | Details on cancers diagnosed in Scotland | Tumour type, morphology, date of diagnosis | 04.2004 – 05.2024 |
| SRR [18] | Details on patients with kidney disease | Kidney disease diagnoses, date/type of treatments, transplant records | 04.2004 – 09.2025 |
| SCI Store [19] | Laboratory investigations – biochemistry and haematology | Test name, test date, test value | To be defined on project level |
| ECOSS [20] | Laboratory investigations – microbiology | Microbiological test results, specimen, organism, sensitivity, MIC | 01.2019 – 12.2025 |
| **Hospitalisations** |  |  |  |
| SMR00 [21] | Outpatient clinic attendances | Clinic speciality, referral reason, date of clinic attendance | 04.2004 – 06.2025 |
| SMR01 [22] | Hospital inpatient and day-case episodes | Admission date, admission reason (ICD-10), surgical procedure (OPCS-4), discharge date, length of inpatient stay | 04.2004 – 06.2025 |
| SMR02 [23] | Maternity Care | Date of delivery, admission/discharge date, admission reason (in labour/after delivery at home), diabetes(Y/N) | 04.2004 – 06.2025 |
| SMR04 [24] | Mental health inpatient and day-case admissions | Admission/discharge date, reason for admission, admission type (emergency/routine) | 04.2004 – 06.2025 |
| SICSAG [12] | Intensive care unit (ICU) admissions | ICU admission date, organ support, interventions | 01.2005 – 12.2024 |
| **Mortality** |  |  |  |
| NRS Death Records [25] | Civic registration system | Date of death, cause of death | 04.2009 – 10.2025 |
| **Patient demographics** |  |  |  |
| CHI |  | Age, sex |  |
| various | Geographical information based on postcode | Health board of residence, urban-rural classification, deprivation (SIMD) |  |

CHI – Community Health Index; ECOSS – Electronic Communication of Surveillance Scotland; HCM – Home Care Medicines; HEPMA – Hospital Electronic Prescribing and Medicines Administration; NRS – National Records of Scotland; PIS – Prescribing Information System; SCI – Scottish Care Information; SICSAG – Scottish Intensive Care Society Audit Group; SIMD – Scottish Index of Multiple Deprivation; SMR – Scottish Morbidity Records; SRR – Scottish Renal Registry. In HCM, supply date corresponds to delivery date.

Source extracts from PHS datasets are ingested into a controlled staging layer in which original dataset structures are preserved, and initial data quality checks are performed (Figure 1). Within the National Safe Haven (NSH), a secure environment used to enable researcher access to pseudonymised individual-level data, MACCS is organised using a structured database design (or schema) that defines how different types of information are stored, related, and queried. The core schema includes person-level tables that hold basic demographic information for each individual, alongside harmonised medicines exposure tables derived from the PIS, HEPMA and HCM, which record medicines prescribing and supply (where available) in a consistent format. Additional event-based tables capture linked information on hospital activity, laboratory results, disease registries, and mortality data. Data is updated at regular intervals through repeat data extractions from the source datasets; this approach preserves a consistent data structure while allowing new records to be incorporated over time. All records include dates of relevant events, allowing researchers to follow individuals over time, subject to data availability, and to support longitudinal analyses, the use of look-back periods, and time-to-event study designs.

**Figure 1.**
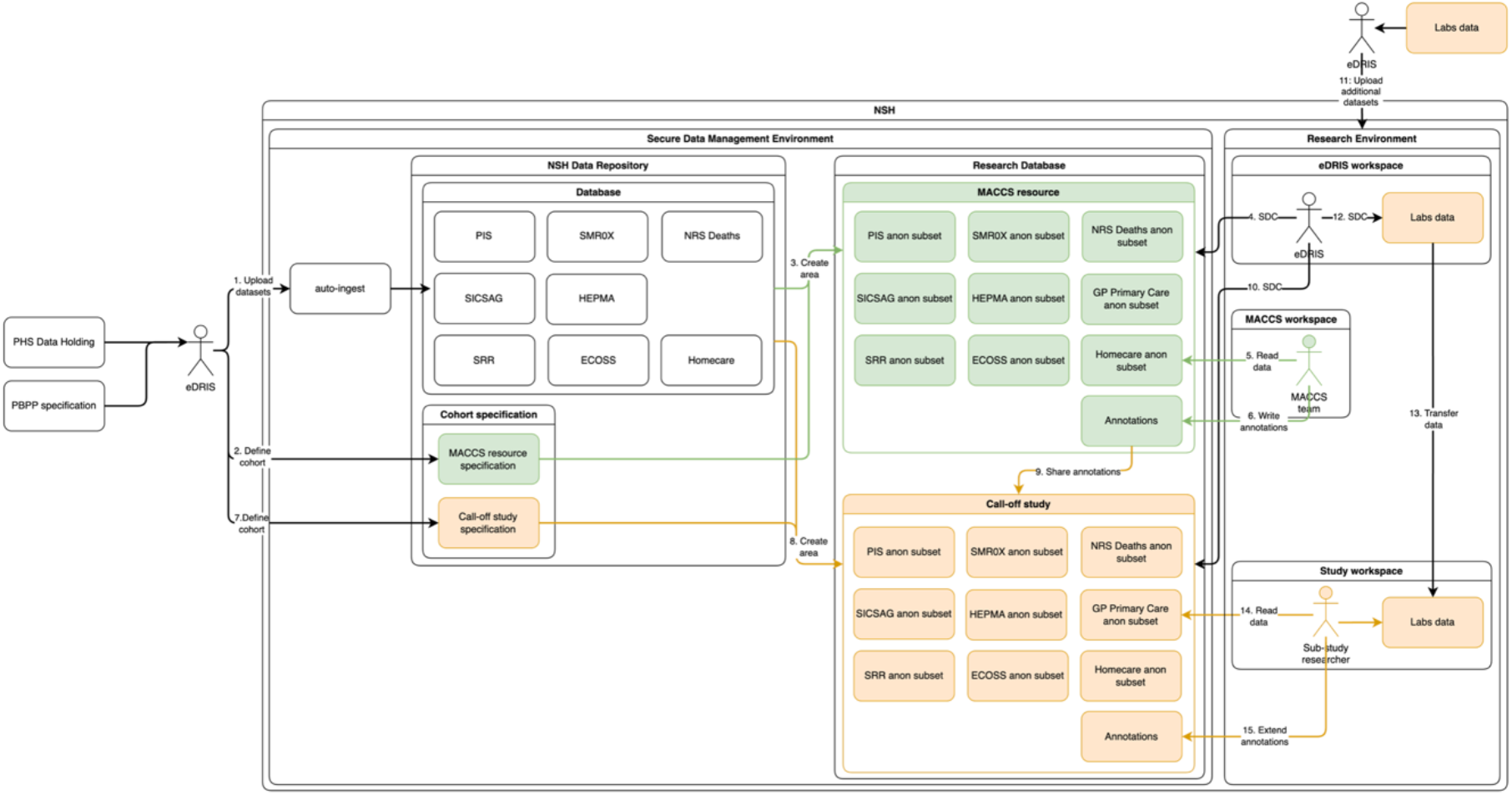
Medicines in Acute and Chronic Care in Scotland (MACCS) architecture schematic. This schematic illustrates the operation of the MACCS data resource. Source datasets are held within the National Safe Haven secure data environment and are managed by the eDRIS team through the NHS Data Repository Database, alongside MACCS project specification code. To create the MACCS research environment, national datasets are linked, cleaned, and standardised in line with MACCS resource specifications. For each approved study, project-specific data views are generated from the core resource based on defined call-off requirements and made available to researchers through controlled analytical workspaces with integrated governance and auditing.

Variables are harmonised across datasets as far as possible (e.g., consistent variable naming and formatting); standardised coding systems are used where available and appropriate. These include, e.g., British National Formulary (BNF) coding [29] or medicines dispensed in community pharmacies and Dictionary of Medicines & Devices (dm+d) codes [30] for medicines prescribed and administered in hospital settings, both of which can be mapped to the Anatomical Therapeutical Chemical (ATC) classification system [12]; and International Classification of Diseases 10^th^ Revision (ICD-10) codes [31] to capture reasons for admission to hospital and causes of death. Dates are recorded using standard ISO formats, and categorical variables use clearly defined code lists.

## Data resource use

MACCS can support a wide range of projects in areas such as pharmacoepidemiology, pharmacovigilance, and health services research due to the extent of data available. This may include the evaluation of prescribing practices across Scotland’s care settings and the tracking of real-world medicines use on a population level; the assessment of patient adherence to medication; long-term safety monitoring of medication use; or assessing associations between medication use, laboratory markers, and clinical outcomes over time. Furthermore, MACCS and the infrastructure it sits in can support various methodological approaches, including advanced statistical methods and application of Artificial Intelligence (AI) tools.

An illustrative example of the current application of MACCS is a study examining high-dose antipsychotic prescribing in hospitalised patients. The aim of the study is to assess current prescribing practices and evaluate the association between high-dose antipsychotic prescribing and clinical outcomes among hospitalised patients; more specifically, objectives include calculating the prevalence of high-dose antipsychotic prescribing across different hospital settings; describing patient characteristics and clinical profiles of patients exposed to high-dose antipsychotic therapy; and analysing both short- and long-term effects of high-dose antipsychotic use (e.g., adverse events, subsequent hospitalisations, and mortality). MACCS facilitates the identification and characterisation of prescribing patterns and patient exposure across care settings, both in primary and secondary care; moreover, MACCS provides access to crucial laboratory data as well as the required data on hospitalisations and death. Findings from this study will be used to inform the development of a subsequent project focused on designing an integrated computerised clinical decision support (CDS) tool to support safer antipsychotic prescribing in clinical practice. Further details can be found in the protocol [32].

MACCS is also currently being used to support a feasibility study evaluating real-world safety surveillance of targeted immune-modifying therapies across immune-mediated inflammatory disease in Scotland, commissioned by the Medicines and Healthcare products Regulatory Agency (MHRA). By linking HCM data with hospital admissions, the cancer registry, and death records, MACCS enables the identification of new users of biologics and Janus kinase (JAK) inhibitors and the estimation of incidence rates for serious infections, major cardiovascular events, venous thromboembolisms, and malignancies. The work demonstrates the capacity of MACCS to underpin scalable, population-level pharmacovigilance within a national regulatory context.

Additional studies using MACCS are in various stages of planning. For instance, a study examining anticoagulant prescribing patterns over time and associated clinical outcomes has recently been approved; this study aims to develop and validate a prediction model to estimate the risk of severe bleeding in patients treated with Direct Oral Anticoagulants (DOACs) in Scotland. The study will quantify how often serious bleeding events (hospitalisation or death) occur and identify key demographic, clinical and treatment-related risk factors. Different statistical approaches will be compared to determine which most accurately predicts bleeding outcomes. Using MACCS resource, the study aims to create a robust multivariable model that estimates individual risk at treatment initiation, supporting clinicians to identify high-risk patients and guide safer, personalised prescribing and monitoring decisions. Another planned study will focus on the longterm effects of COVID-19 infections in the Scottish population, exploring medication use for psychiatric disorders following infection, describing treatment pathways, and assessing associated clinical outcomes.

## Strength and weaknesses

A major strength of MACCS is its population-scale coverage within a universal healthcare system, enabling near-complete ascertainment of medicines prescribing, dispensing, administration, and/or supply for adults receiving NHS Scotland care. Because data are captured routinely whenever individuals interact with NHS services, which are provided free at the point of care to all residents, MACCS supports longitudinal analyses without the need for sampling and therefore minimises selection bias commonly encountered in cohort studies. The integration of medicines data from different settings within a single, pre-linked resource allows comprehensive assessment of medicines use and treatment pathways across NHS Scotland care. A further strength is the linkage of medicines exposure data to a wide range of national clinical datasets, including hospital admissions, laboratory results, and mortality records. This enables robust investigation of treatment outcomes, adverse events, and safety signals in real-world settings. The provision of data within MACCS through curated schema and harmonised variables reduces the technical burden associated with assembling multi-source datasets and supports reproducible research through consistent data structures and periodic refreshes. Finally, MACCS also benefits from a streamlined governance and access model. Approved researchers only have to apply once to access a pre-linked data resource within a secure Trusted Research Environment, rather than having to navigate multiple dataset-specific approvals. This improves efficiency while maintaining robust information governance, privacy protection, and disclosure control.

Limitations of MACCS mainly reflect those inherent to routinely collected administrative data. First, some desirable information may potentially be incomplete; for example, the clinical indication for prescribing is currently not available within PIS or HEPMA, thus linkage to other datasets (e.g., hospital admissions) may be required. Second, data is not entirely standardised due to differences in how – and for what original purpose – data in the underlying individual datasets was collected; in HCM for instance, the date of prescription is approximated by the date of delivery, which may affect the accuracy of analyses specifically focusing on prescribing intent. And third, although data is updated on a regular basis, there is a time lag to allow for routine quality checks prior to making data available to researchers, with potential implications for time-sensitive research projects. In addition, MACCS only provides information on adults. Despite these limitations, MACCS represents the most comprehensive resource currently available for population-scale evaluation of medicines use and associated outcomes in Scotland, and enables longitudinal, population-scale studies that may inform policy, clinical guidelines, and the optimisation of patient care.

## Data resource access

MACCS is not an open-access dataset due to the inclusion of person-level health data. Consequently, no public web-based download is available. Access to the data is granted only for approved research projects and is provided exclusively within secure Trusted Research Environments in accordance with NHS Scotland information governance requirements. Researchers seeking access to MACCS for research purposes must submit an application through eDRIS at Public Health Scotland; all requests are subject to review and approval by PBPP-HSC [13].

Supporting documentation for MACCS, including variable lists and data dictionaries describing variable definitions, derivation rules, and coding systems, is available to approved users. These materials are provided as part of the data access process and are maintained by PHS. Variables are presented in a structured, analysis-ready format, with consistent naming conventions applied across data sources; variable names typically follow clear, descriptive rules, using standardised prefixes and suffixes to distinguish patient-level characteristics, medicines exposure variables, and derived measures. No specialist software is required to access MACCS beyond what is provided within the Trusted Research Environment. Approved users can analyse the data using standard analytical tools such as R and Python; analyses can be conducted remotely, although researchers need to be UK-based to be able to access the remote server. Nevertheless, international collaboration is encouraged. Researchers outside the UK are invited to collaborate with UK-based academic or NHS partners who are able to host and conduct analyses within the secure infrastructure. Such collaborations support wider national and international research while ensuring compliance with data protection and confidentiality requirements.

Enquiries regarding the MACCS data resource, including questions about data content, access processes, and potential collaboration, can be directed to the corresponding author: Cosmika Goswami, University of Strathclyde; or to PHS directly on.

## Data Availability

All data produced in the present study are available upon reasonable request to the authors

## Ethical approval

The development and setting up of MACCS was approved through the Public Benefit and Privacy Panel for Health and Social Care. Additional ethical approval was not required. No data was collected or generated specifically for this project.

## Author contributions

All authors substantially contributed to the conception and design of the work. CG and TM drafted the initial manuscript, with input from all authors. All authors critically reviewed and revised the draft; and approved the final manuscript.

## Acknowledgements

We thank NHS Scotland and Public Health Scotland (PHS) for their support in facilitating access to the datasets. We are also grateful to the healthcare professionals and clinicians whose input has helped shape the scope and content of the data resource.

